# Low serum levels of magnesium, phosphate, and zinc in ICU patients – prevalence, management, characteristics, and outcomes Protocol and statistical analysis plan for The WhyTrace cohort study

**DOI:** 10.1101/2022.05.11.22274933

**Authors:** Gitte Kingo Vesterlund, Hans-Christian Thorsen-Meyer, Benjamin Skov Kaas-Hansen, Morten Hylander Møller, Anders Perner

## Abstract

**Background:** Patients admitted to the intensive care unit often have low serum levels of magnesium, phosphate, and zinc. We aim to describe the prevalence, management, characteristics, and outcomes of patients with low levels of these minerals in a large cohort of Danish ICU patients.

**Methods:** In this multicenter cohort, we will include all adult ICU patients admitted to one of 10 ICUs during a six years period. Patients will be excluded if the ICU admission is post-operative after planned surgery, length of stay is <24 hours, they are transferred from ICUs not participating in the study, or are without measurements of neither magnesium, phosphate nor zinc. First, we will do a descriptive assessment of the prevalence of low serum levels and management of supplementation according to different patient characteristics. Second, we will conduct a cohort study using prospectively registered outcome data, to assess associations between serum levels of magnesium, phosphate, and zinc respectively, and each of two principal outcomes: time to successful extubation and incident tachyarrhythmia. We will use a joint modelling approach for estimating the strengths of associations, combining time to event data and longitudinal data from repeated measurements of serum levels.

**Discussion:** This study will provide information from a very large cohort of Danish ICU patients. Strengths of the study will be the large number of patients, the interconnection of data from several registers, and for the association analysis the use of a statistical approach making the optimal use of the repeated measures of serum levels. Limitations will be the risk of bias from unmeasured or unregistered confounding factors, and the lack of randomization in the observational design, which will prevent us from interpreting possible associations between serum levels and outcomes as definite effects on the outcome.

## 1. Introduction

### 1.1 Background

Patients admitted to the intensive care unit (ICU) often have low serum levels of magnesium, phosphate, and zinc. ^1-3^ This may partly be caused by large fluid shifts and redistribution during the ICU stay. ^2,4,5^ However, low serum levels have been reported even at baseline when patients are admitted to the ICU.^2,4,6^ We have previously found a marked variation in the reported assessment and administration of magnesium, phosphate and zinc among ICUs.^7^ As an important next step, we aim to describe the clinical landscape including the prevalence, management of supplementation, characteristics and outcomes of patients with low levels of these minerals in a large cohort of Danish ICU patients.

### 1.2 Objectives

Our aims are

- To assess the prevalence of low serum levels of magnesium, phosphate, and zinc: how many patients have low, high, and normal serum levels at baseline, and how many develop it during their ICU stay?
- To assess what characterizes patients with low and high serum levels compared to those with normal serum levels, and what characterizes patients to whom supplementation therapy is given once or more than once.
- To assess whether deviating serum levels of magnesium, phosphate or zinc are associated with time to successful extubation and/or mortality, and if low serum levels of magnesium are associated with incident tachyarrhythmia.

We expect that hypomagnesemia, hypophosphatemia and hypozincemia are frequent and expect that there may be substantial variation in the rates of supplementation of these trace elements among patients with different characteristics in the ICU.

We further hypothesize that hypomagnesemia, hypophosphatemia and hypozincemia may be associated with increased time to successful extubation, and hypomagnesemia with incident tachyarrhythmia.

## 2. Methods

### 2.1 Study design

In this multicenter cohort of Danish ICU patients we will first, do a descriptive assessment of serum levels and supplementation and, second, gauge the associations between low serum levels and patient outcomes using prospective data. Findings will be presented in accordance with the STrengthening the Reporting of Observational studies in Epidemiology’ (STROBE) statement^8^ and the protocol will be publicized online at medRxiv.org before data processing and analysis begin.

### 2.2 Study population

#### Inclusion criteria

All adult patients admitted to one of 10 general ICUs in Denmark between October 2011 and January 2018. We expect to include thousands of patients and so power calculations are foregone due to the expectedly large number of patients.

#### Exclusion criteria

- Post-operative ICU admission after planned surgery
- Length of ICU stay <24 hours
- Patients transferred from ICUs not participating in the study (due to missing baseline measures)
- Patients without serum measurements of neither magnesium, phosphate nor zinc

### 2.3 Variables

Exposure variables:

- Baseline serum levels of total magnesium, phosphate, and zinc at ICU admission (defined as a sample drawn between 24 hours prior to ICU admission and 24 hours after ICU admission. If more than one sample is available, we will use the lowest value)
- All subsequent serum levels of total magnesium, phosphate and zinc during ICU including timestamp. For the descriptive analyses, serum levels of magnesium, phosphate and zinc will be grouped into low, normal, and high serum levels as follows. Magnesium: <0.70, [0.70-0.95], >0.95 mmol/L. Phosphate: Females: <0.76, [0.76-1.41], >1.41 mmol/L. Males age <50 years: <0.71, [0.71-1.53], >1.53 mmol/L. Males age >50 years: <0.71, [0.71-1.23], >1.23 mmol/L. Zinc: <10, [10-19], >19 μ mol/L.^9^ For the association analyses, actual serum levels will be used.
- Daily administration of magnesium, phosphate and zinc either orally or intravenously will be registered as yes/no

Patient characteristics:

- Age at inclusion
- Sex
- Severity of illness as SOFA (Sepsis-related Organ Failure Assessment) score^10^ (0-24) at baseline
- Arterial pH (defined as lowest value within 24 hours of ICU admission)
- Medical vs. surgical patient category
- Malignancy (defined as metastatic cancer or hematologic cancer disease, either in patient record at ICU admission or with ICD-10 codes DC 77-79 and DC 81-95 in NPR from 1994 and prior to current admission)
- COPD (chronic obstructive pulmonary disease) or asthma (either in patient record at ICU admission, or with ICD-10 codes DJ40-47 in NPR from 1994 and prior to current admission
- Heart disease (defined as previous acute myocardial infarction or chronic heart failure either in patient record at ICU admission, or with ICD-10 codes DI20, 22, 50, 252, 110, 130 or 132 in NPR from 1994 and prior to current admission)
- Severe kidney-failure (defined as either CRRT or hemodialysis during first 24 hours of ICU stay or chronic dialysis, either in patient record at ICU admission or in NPR)
- Liver cirrhosis (either in patient record at ICU admission, or ICD-10 codes DK703, 703A, 717, 732E or 74 in NPR from 1994 and prior to current admission)

The patient characteristics are based on previous evidence suggesting the variable to be a predictor of the outcome, or on clinical experience indicating a variable to be so despite lack of studies examining an association, when the variable previously has been associated with the trace-elements phosphate, zinc or magnesium distribution.

### 2.4 Descriptive statistics

Categorical variables will be reported as count (percentage) and continuous values as median (inter-quartile range (IQR)). There will be nine baseline serum level groups: hypo-, normo-, and hypermagnesemia, -phosphatemia, and -zincemia.

The following descriptive statistics will be reported in the manner described:

- Number of included patients in each baseline serum level group
- Overall prevalence of hypo- and hypermagnesemia, -phosphatemia, and -zincemia during ICU will be assessed by using each patient’s lowest and highest serum values.
- Timing of hypomagnesemia, -phosphatemia, and -zincemia will be presented in histograms (cumulative if more informative) of the number of patients with incident low serum levels at days 0, 1, 2, 3 through 28 and >28 days.
- Prevalence and duration of oral or intravenous magnesium, phosphate, or zinc respectively during ICU. Duration will be reported as proportion of ICU days with supplementation.
- To describe what characterises patients with low versus normal and high versus normal baseline serum levels of magnesium, phosphate, and zinc, we will report patient characteristics stratified by baseline serum level group.
- To describe what characterises patients who receive supplementation with magnesium, phosphate and zinc, patient characteristics will be stratified by groups: Supplementation therapy once during ICU, supplementation therapy several times during ICU and no supplementation received of magnesium, phosphate and zinc (9 groups in total).
- Time to successful extubation in patients alive after 28 days stratified by baseline serum level groups and the proportion of patients with tachyarrhythmia onset in each of the magnesium groups.
- 28-days mortality by baseline serum level group.

Patients who are transferred to an ICU which are not participating in the study will not be included in the descriptive assessments.

### 2.5 Statistical analysis of associations between serum levels and patient outcomes

In the analytical part, we will conduct a cohort study using prospectively registered outcome data, to assess associations between serum levels of magnesium, phosphate, and zinc respectively, and each of two principal outcomes (detailed below).

#### Outcomes

- Time to successful extubation. To avoid bias from unsuccessful extubations resulting in reintubation, we will count the days from first intubation until final extubation (i.e. if a patient is extubated on day four, reintubated on day six and finally extubated on day 7, the count will include the full seven days).^11^ Follow-up will continue if patients are discharged to a non-ICU department.
- Incident tachyarrhythmia (defined as a first heartrate >120 during ICU, where clinicians subsequently administered intravenous amiodarone or digoxin). Patients discharged to a non-ICU department without the event will be considered censored. This outcome will only be analyzed in the magnesium exposure model.

Follow-up will be capped at 28 days. When patients are transferred to an ICU not participating in the study, they will be censored due to missing information on the outcome.

#### Covariates

The patient characteristics listed above will be used as covariates in the models.

#### Statistical models

To make full use of the longitudinal data on serum levels of magnesium, phosphate and zinc, we will use a joint modelling approach for estimating the strengths of associations between serum levels and the two outcomes (time to successful extubation and incident tachyarrhythmia). In essence, a joint model combines two types of sub-models to yield combined effect size estimates in the unit of hazard ratios.^12-14^ The first type (called the event sub-model) uses time to event data to conduct a survival analysis. Mortality will be considered a competing risk for both outcomes and discharge from the ICU a second competing risk for incident tachyarrhythmia. The event sub-model uses the baseline covariates for adjustment.

The second type of sub-model (the longitudinal sub-model) uses data from repeated measurements of (in our case) serum levels and usually takes the form of a linear mixed-effects model.

A crucial component is the association structure between the longitudinal sub-model and the event sub-model. We will follow the most common tactic and use the expected or predicted value of the longitudinal sub-model at the time to event/censoring. However, since we anticipate a U- or J-shaped effect of serum levels (both low and high values are harmful), we will use linear splines in the association structure to allow such non-linear effects to emerge.^15^ Due to the expected large number of patients without serum-level data in all three trace elements, we will build and fit one model per trace element per outcome.

#### Readmission to the ICU

Each patient can only be included once. In case of multiple ICU admissions, we will use the first. Admission to another ICU within 6 hours from discharge will be considered a transfer and therefore a continued admission.

#### Missing data and subgroup analysis

To discuss potential differences between included and excluded patients we will conduct univariate comparisons of the baseline characteristics between the groups. Likewise, we will compare patients with missing baseline measures of magnesium, phosphate or zinc to patients with baseline measures. In case of ≤5% missing observations on other variables we will perform the analyses in the complete case dataset. If >5% observations are missing, we will consider using imputations. In that case, the primary results will be based on these data and complete case analyses will be presented in a supplementary file.

### 2.6 Data sources

Information on duration of mechanical ventilation, incident tachyarrhythmia, age, sex, SOFA score, medical vs. surgical patient category, dialysis first 24 hours, arterial pH and supplementation therapy will be obtained from the electronic patient record system CIS (Cambio Healthcare Systems A/S, Copenhagen) containing patient medical records. Information on transfers and co-morbidities will be obtained from the Danish National Patient Register (NPR), containing information on e.g. admissions and diagnoses since 1977.^16^ To reduce the risk of misclassification from possibly missing ICD10 codes in NPR, co-morbidities will also be collected if registered in CIS at ICU admission. Laboratory measurements (serum magnesium, phosphate, and zinc) will be obtained from the electronic laboratory database LABKA II or BCC containing all test results for the Capital Region and Region of Southern Denmark, respectively. Information from all sources will be combined using the personal identification number which is assigned to all permanent Danish residents and registered in the Danish Civil Registration System (CRS),^17^ from where mortality also will be obtained.

### 2.7 Data and code availability

The data we will assess for this study cannot readily be shared because of national and EU regulations. However, data is available for use in secure, dedicated environments via application to the Danish Patient Safety Authority and the Danish Health Data Authority. Analytical code used in the study will be made publicly available.

## 3 Discussion

This study will provide information on prevalence of low serum levels of magnesium, phosphate and zinc, the management of supplementation, patients characteristics associated with supplementation and low serum levels, from a very large cohort of Danish ICU patients. Furthermore, it will provide estimates of associations between serum levels and patient outcomes. The strengths of the study include the large number of patients, from multiple centers, the use of the unique Danish personal identification number enabling interconnection of several registers with minimal loss to follow-up, and for the association analysis the use of a statistical approach making the optimal use of the repeated measures of serum levels. Limitations will be the risk of bias from unmeasured or unregistered confounding factors, and the lack of randomization in the observational design, which will prevent us from interpreting possible associations between serum levels and outcomes as definite effects on the outcome.

## 4 Ethical considerations

The study has been approved by the Danish Data Protection Agency (DPPA) and the Danish Patient Safety Authority (DPSA). All data will be handled with confidentiality and participants will be anonymized.

## Acknowledgements

The study was funded by the Research foundation of Rigshospitalet, Copenhagen University Hospital, Denmark; Novo Nordisk Foundation (Grant agreement NNF14CC0001) and the Innovation Fund Denmark (Grant agreement 5153-00002B).

